# A Reagent and Virus Benchmarking Panel for a Uniform Analytical Performance Assessment of N Antigen–Based Diagnostic Tests for COVID-19

**DOI:** 10.1101/2022.08.03.22278351

**Authors:** Allison Golden, Jason L. Cantera, Lorraine Lillis, Thanh T. Phan, Hannah Slater, Edwin J. Webb, Roger B. Peck, Gonzalo J. Domingo, David S. Boyle

## Abstract

Rapid diagnostic tests (RDTs) that detect antigen indicative of SARS-CoV-2 infection can help in making quick health care decisions and regularly monitoring groups at risk of infection. With many RDT products entering the market, it is important to rapidly evaluate their relative performance. Comparison of clinical evaluation study results is challenged by protocol design variations and study populations. Laboratory assays were developed to quantify nucleocapsid (N) and spike (S) SARS-CoV-2 antigens. Quantification of the two antigens in nasal eluates confirmed higher abundance of N than S antigen. The median concentration of N antigen was 10 times greater than S per genome equivalent. The N antigen assay was used in combination with quantitative RT-PCR to qualify a panel composed of recombinant antigens, inactivated virus, and clinical specimen pools. This benchmarking panel was applied to evaluate the analytical performance of the SD Biosensor STANDARD Q COVID-19 Ag test, Abbott Panbio COVID-19 Ag Rapid Test, Abbott BinaxNOW COVID-19 Ag test, and the LumiraDx SARS-CoV-2 Ag Test. The four tests displayed different sensitivities toward the different panel members, but all performed best with the clinical specimen pool. The concentration for a 90% probability of detection across the four tests ranged from 21 pg/mL to 102 pg/mL of N antigen in the extracted sample. Benchmarking panels provide a quick way to verify the baseline performance of a diagnostic and enable direct comparison between diagnostic tests.

## INTRODUCTION

Diagnostic tools are essential for surveillance and control of the COVID-19 pandemic.^1^ While reverse transcription polymerase chain reaction (RT-PCR) from a nasopharyngeal or nasal swab is the gold standard for confirmation of infection with SARS-CoV-2, the complexity of such tests requires sophisticated laboratory systems, and imposes logistical challenges for its effective use in scenarios requiring either a fast time-to-result or where laboratory systems are not robust. Rapid diagnostic tests (RDTs) designed to detect viral antigens, primarily the nucleocapsid protein (N) antigen, hold promise for testing in settings where RT-PCR cannot be implemented, and as both time and cost-saving measures for frequent testing and entry points.^1–3^

The earliest-to-market RDTs were tested in clinical studies to assess their performance and utility. The results of these clinical studies are informative in terms of clinical performance within the context of the studies conducted, but they also highlight the variability in clinical performance as driven by the study design, target population, and other study-specific factors. ^4–6^

There is need for a performance assessment of these tests that is less study specific and can be performed in multiple laboratories, the results of which would enable more direct comparison of performance across different RDTs.^7^ If the results of this assessment can be linked to clinical data, they may also be indicative of clinical performance. Currently, assessments of analytical performance have been expressed primarily through comparison to SARS-CoV-2 viral RNA in terms of cycle thresholds (Ct) or to cultured virus infective units, with increasing correlation to viral copy number quantification to aid in standardization of results.^7–13^ Complementary to these efforts, this article presents a quantitative open-platform assay for the N and spike (S) antigens, a comparison of genome equivalents (GEs) to N and S antigen concentration from clinical samples, a panel of reagents with which to assess the performance of the RDTs representing multiple sources of target analyte protein, and the results from assessment of four different emergency use–authorized (EUA)/licensed (EUL) COVID-19 rapid antigen diagnostic tests.

## MATERIALS AND METHODS

### Materials

Full-length recombinant N antigens expressed in *Escherichia coli* and in HEK293 mammalian cells were purchased from Native Antigen (Kidlington, United Kingdom) and Acro Biosystems (Newark, Delaware, USA), respectively. Recombinant S antigen, in a stabilized trimeric form and expressed in HEK293 mammalian cells, was purchased from Acro Biosystems.

The following reagents were obtained through BEI Resources (Manassas, Virginia, USA); and the US National Institute of Allergy and Infectious Diseases (NIAID)/US National Institutes of Health (NIH), contributed by the US Centers for Disease Control and Prevention (CDC): SARS-Related Coronavirus 2, Isolate USA-WA1/2020, Gamma-Irradiated (NR-52287), and Genomic RNA from SARS-Related Coronavirus 2, Isolate USA-WA1/2020 (NR-52285). The following reagents were obtained through BEI Resources and NIAID/NIH: Human Coronavirus 229E (NR-52726), and Human Coronavirus OC43 (NR-52725). NR-52287 used for N antigen determination had assigned values for the concentration of infectious virus of 2.8□×□10^5^ 50% tissue culture infective dose (TCID_50_)/mL and RNA (4.1□×□10^9^ copies/mL), determined prior to inactivation. The viral RNA control for RT-PCR was prepared from USA-WA1/2020 (47.5 ng/mL total RNA with an estimated 1.84 × 10^7^ GEs/mL).

Using the GenBank sequence of the 419 as a full-length SARS-CoV-2 N protein, GenBank accession number QHO62115.1, with an additional cleavage site and a polyhistidine tag, an extinction coefficient was calculated based on ProtParam^14^ of Abs 0.1% (= 1 g/L) of 0.959. This extinction coefficient was used to confirm stock protein concentration of recombinant SARS-CoV-2 antigen that was aliquoted and stored at −80°C.

The buffer diluent contained 1X phosphate-buffered saline (10 mM PBS, 2.7 mM potassium chloride, 137 mM sodium chloride pH 7.4) with 1% bovine serum albumin (BSA), 1X PBS + 1% BSA. Negative swab pool diluent contains pooled discarded SARS-CoV-2 PCR-negative human nasal swabs eluted into 1X PBS.

### Clinical samples

De-identified nasal swab eluates were acquired from the Washington COVID-19 Biorepository (Seattle, Washington, USA) or Boca Biolistics (Pompano Beach, Florida, USA). Nasal swab eluates used in this study were prepared in either 1X PBS or Clinical Transport Medium (Noble Biosciences, Gyeonggi-do, Republic of Korea).

### Quantification of SARS-CoV-2 N and S antigens using SARS-CoV-2 antigen immunoassays

An immunoassay detecting SARS-CoV-2 N and S antigens was developed using the Meso Scale Discovery (MSD) platform (Meso Scale Diagnostics, Rockville, Maryland, USA), which uses electrochemiluminescence for detection.

Antibodies sourced from Sino Biological (Beijing, P.R. China) was used for N antigen detection and an antibody pair from Leinco Technologies (Fenton, Missouri, USA) for S antigen detection.

The capture antibodies were labeled with biotin using the EZ-Link^®^ Sulfo-NHS-LC-LC-Biotinylation kit (ThermoFisher Scientific, Waltham, Massachusetts, USA) and the detector antibodies were labeled with SULFO-TAG™ (GOLD SULFO-TAG NHS-Ester, Meso Scale Diagnostics). Any unbound biotin or SULFO-TAG was removed using desalting columns (Zeba™, 40k MWCO, ThermoFisher Scientific). The concentrations of antibodies were measured at 280 nm via a spectrophotometer (NanoDrop™ 2000C, ThermoFisher Scientific), and concentration of detector antibody following labeling was assigned 90% of the concentration prior to desalting. Standards were prepared from recombinant HEK293-expressed full-length SARS-CoV-2 N protein and stabilized trimeric S protein (Acro Biosystems).

### Clinical sample testing with SARS-CoV-2 antigen assays

N and S assays were run separately, using 25 µL per well of the 0.5 µg/mL biotinylated capture antibody was used to coat a blocked SECTOR small spot streptavidin plate (Meso Scale Diagnostics). Analysis of signal and quantification of unknowns relative to the standard curve were conducted using Meso Scale Diagnostics’ Discovery Workbench 4.0 software. For quantification, standards and blank were fit with a 4-parameter log logit fit with 1/y2 weighting. The lower limit of detection (LOD) was defined by the software’s curve fitting. The lower limit of quantification (LLOQ) was defined by the lowest concentration of standard with signal above the following: the limit of blank plus 10 times the standard deviation of the limit of blank.^15^ The upper limit of quantification (ULOQ) was defined by both software and a back-calculated recovery average of 100% ± 20%. Standard curves spanned 0.128 pg/mL to 50 ng/mL of N antigen,^7^ and 0.128 pg/mL to 1,250 pg/mL of S antigen.

The concentration of SARS-CoV-2 N antigen was measured in 405 residual nasal swab eluates, characterized by PCR at CLIA registered clinical laboratories, collected in July through December 2020 in Washington state. Samples were selected across a range of Ct values from original testing, with selection biased toward higher Ct values. Clinical samples either found or anticipated to be over the detection range were diluted either 5-fold or 20-fold to bring them into range, if volume allowed. Replicate well values for positives with a coefficient of variation greater than 20% were repeated.

### Molecular testing for SARS-CoV-2

Viral RNA was extracted from samples using the QIAamp^®^ Viral RNA Mini Kit (Qiagen, Valencia, California, USA) according to the manufacturer’s instructions and eluted in 100 µL buffer. A quantitative RT-PCR (qRT-PCR) assay to estimate the SARS-CoV-2 GE/mL was developed using the N1 primer set developed by the CDC with primers and probe procured from Integrated DNA Technologies (Coralville, Iowa, USA). Each 20 μL final reaction volume contained 5 μL of 4X TaqPath™ 1-Step RT-qPCR Master Mix (ThermoFisher Scientific), 0.5 μL of probe (5 μmol/L), 0.5 μL each of forward and reverse primers (20 μmol/L), 8.5 μL of nuclease-free water, and 5 μL of nucleic acid extract. Amplification was performed on an Applied Biosystems™ 7300 Real-Time PCR instrument (ThermoFisher Scientific). Thermocycling conditions consisted of 15 minutes at 50°C, 2 minutes at 95°C, and 45 cycles of 3 seconds at 95°C and 30 seconds at 55°C. The cutoff for positive samples was less than 40 cycles. The median Cts were used to determine the viral GE concentration using a standard curve from SARS□CoV-2 genomic RNA (Isolate USA□WA1/2020).

### Benchmarking panel

Commercially sourced, full-length, His-tagged recombinant N protein as described above was used. Radiation-inactivated, cultured SARS-CoV-2 virus (BEI Resources, NR-52287) stocks were thawed and diluted into either buffer or negative swab pool. Serial dilutions were further made into diluent and aliquots were frozen. Clinical nasal swab discards from five different individuals, positive for SARS-CoV-2 by qRT-PCR, were selected and combined. The combined samples were then serially diluted into negative swab pool, aliquoted, and frozen. The aliquots were tested by qRT-PCR to quantify viral GE/mL. A benchmarking panel composed of dilutions of recombinant proteins, inactivated viral lysate, and clinical specimen pool was then defined and applied to all diagnostic tests in this study. The panel members were characterized for N antigen concentration using the N antigen immunoassay as well as qRT-PCR for the clinical specimen dilutions. The benchmarking panel is described in Table 1.

**Table 1.**
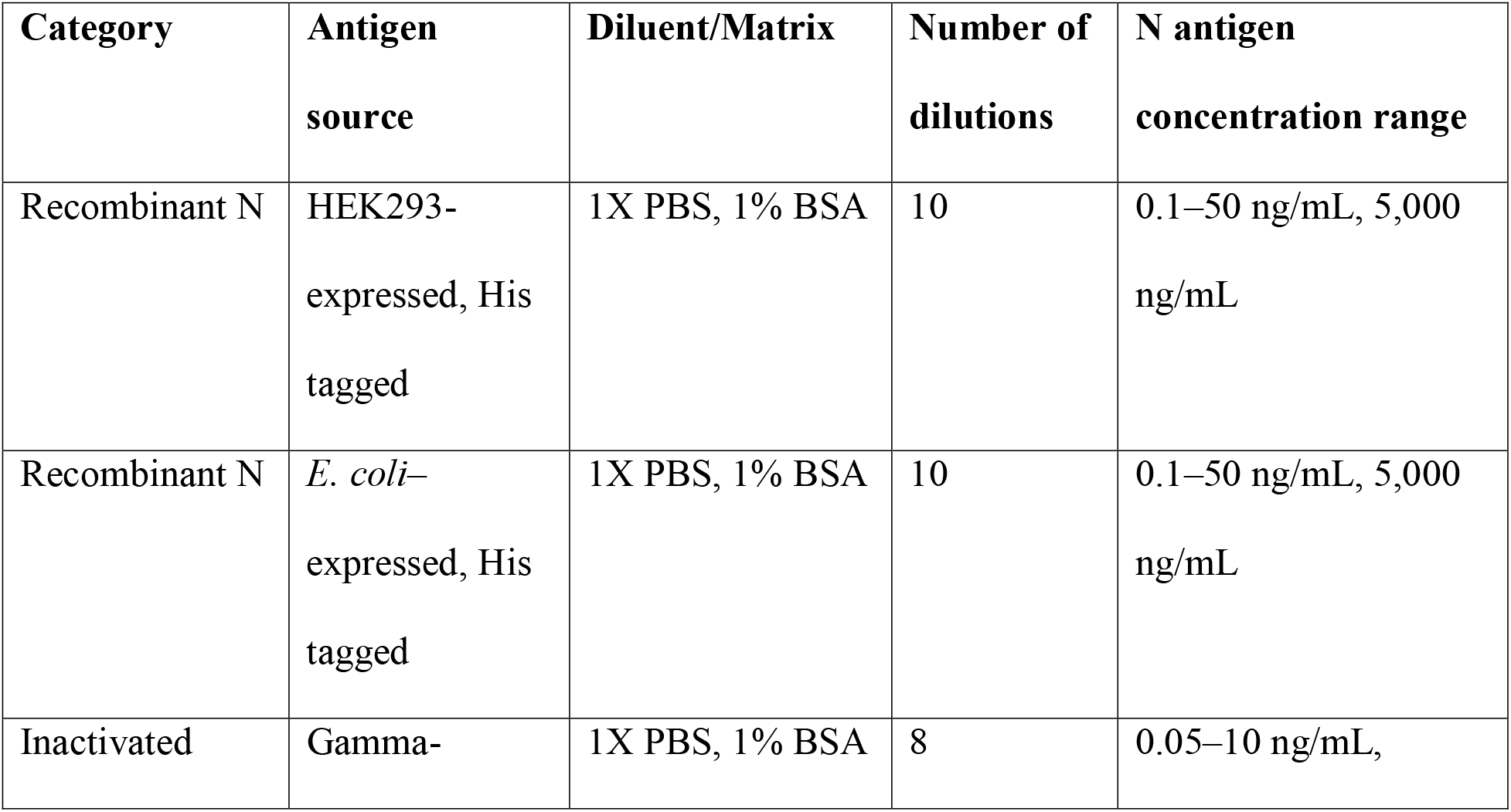
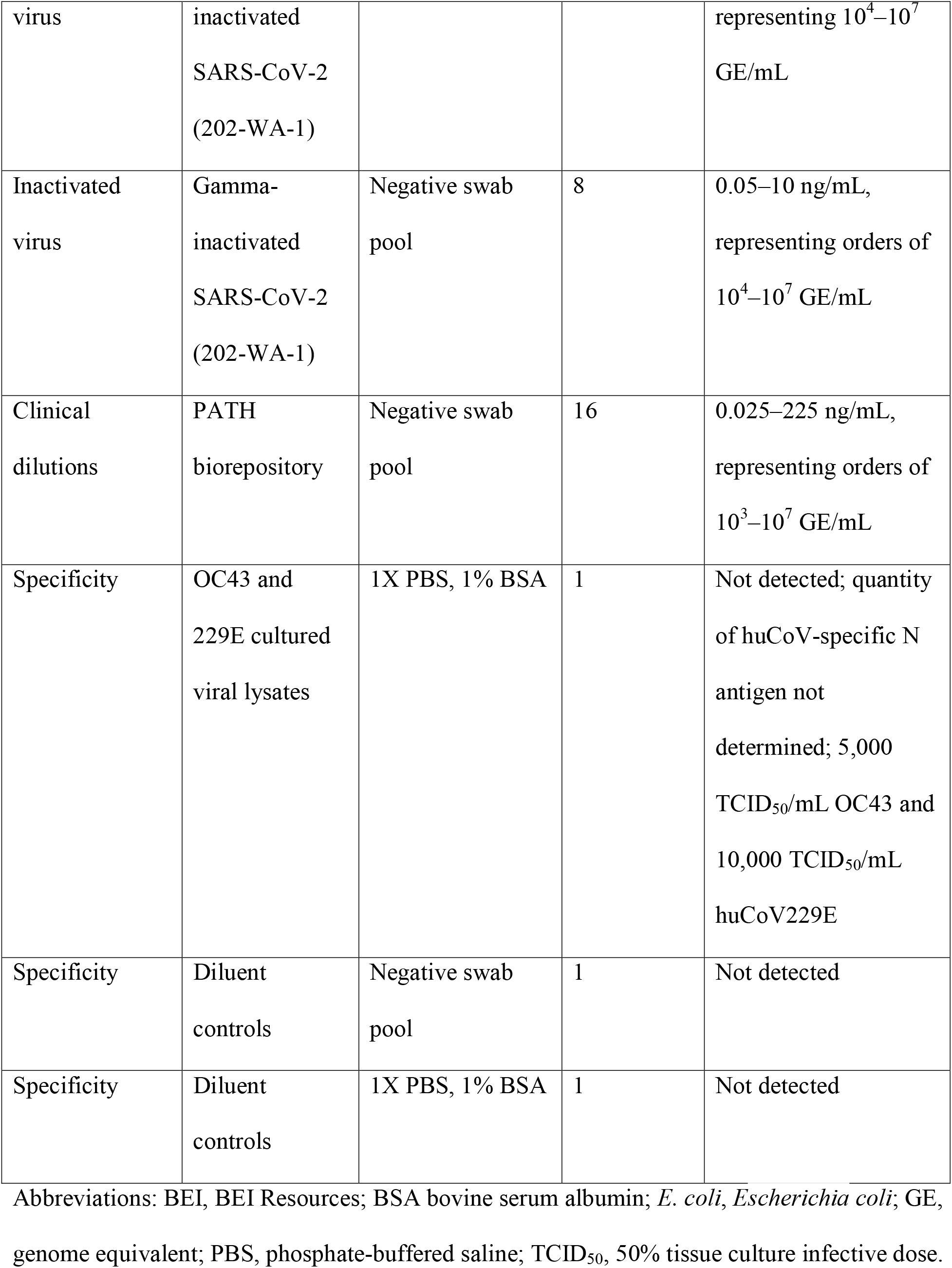
Components of the benchmarking panel used in evaluation of rapid diagnostic tests to detect nucleocapsid and spike SARS-CoV-2 antigens.

### Rapid diagnostic tests used

Four EUA and/or EUL SARS-CoV-2 RDTs were evaluated using N antigen benchmarking panels: the Abbott BinaxNOW™ COVID-19 Ag card test (EUA), Abbott Panbio™ COVID-19 Ag Rapid Test Device (EUL), LumiraDx SARS-CoV-2 Ag Test (EUA), and SD Biosensor STANDARD™ Q COVID-19 Ag Home Test (EUL). Tests were assigned, in no particular order, identification numbers of RDT 1 through RDT 4, for the purpose of this publication, to de-identify specific results.

### Evaluation of rapid diagnostic tests with benchmarking panels

Each RDT was run using a minimum of five replicates per panel concentration and type. Panel member aliquots were thawed on ice and mixed gently. A pipetted volume of the panel mixed into the rapid test–specific extraction buffer simulated the extracted swab material. Thereafter, the instructions for the rapid tests were followed and the diluted panel, at its final concentration in the extraction buffer, was added to the test according to instructions. All panel concentrations were run until two levels of decreasing concentrations were negative for all replicates.

### Statistical analysis of detection limits using benchmarking data

Two statistical models were developed to determine the relationships between (1) analyte concentration and RDT test line intensity and (2) analyte concentration and probability of RDT positivity. For (1), a sigmoid function was fit to the data of the form:

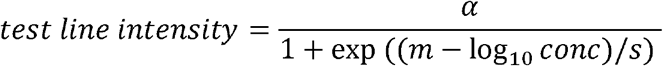

Where *α, m, s* are parameters representing the shape of the sigmoid curve and *conc* is the analyte concentration. This model was separately fitted to each data type (clinical dilution, inactivated virus culture, and recombinant), and the step repeated for all four RDTs. For (2), a simple logistic regression model was fitted to the data, where analyte concentration was the independent variable and the RDT result (1 = positive, 0 = negative) is the dependent variable. A categorical factor representing the data type (clinical dilution, inactivated virus culture, recombinant protein) was included as a covariate to allow for data type–specific fitted curves. This step was repeated for all four RDTs. The models described in (1) and (2) were fitted in a Bayesian framework using the R brms package.^16^ Noninformative Gaussian priors were used for the parameters and the models run for 5,000 iterations after a burn-in of 2,500 iterations. Convergence of chains was assessed using the R-hat statistic and visual checks. The final fitted lines and surrounding shaded areas represent the median and 95% credible of the expected values of the posterior predictive distributions.

### Simulation of detection of clinical samples by rapid diagnostic tests

The detection limits derived from benchmarking data were used to simulate the detection of clinical samples that had been characterized for SARS-CoV-2 N antigen concentration by MSD assay. For each clinical sample, the quantity of N antigen was assumed to be concentrated into the source swab, which had been extracted and diluted into 3 mL transport medium. The quantity of antigen was then divided by the manufacturer-designated volume of rapid test extraction buffer to simulate the final concentration of N antigen that would be added to the test. Finally, this final concentration was compared to detection limits generated in analysis of the benchmarking data to determine whether the sample would be designated as detectable (Figure 1).

**Figure 1.**
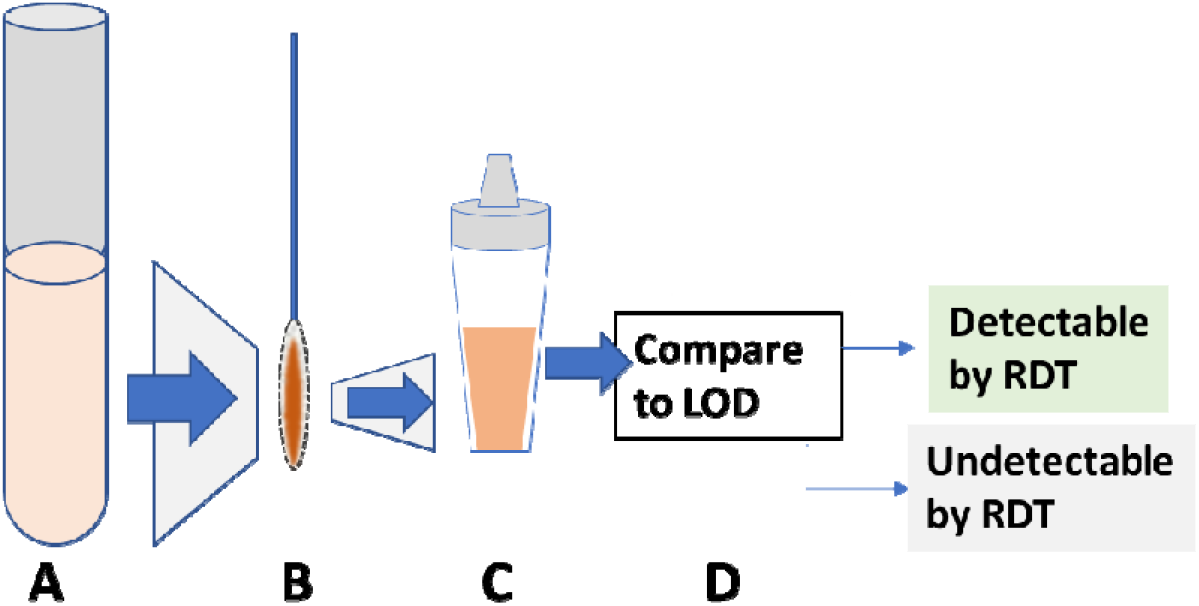
Representation of simulated clinical sample detection by rapid diagnostic test (RDT). **A**. Clinical samples consisting of swab eluate in 3 mL transport medium; N antigen concentration measured. **B**. Total quantity of antigen assumed to be on swab. **C**. Total quantity of antigen is diluted in extraction buffer of RDT to calculate final concentration of N antigen added to RDT. **D**. Final concentration compared to 90% probability lower limit of detection from benchmarking to determine whether detectable by RDT. Abbreviations: LOD, limit of detection; RDT, rapid diagnostic test.

## RESULTS

### Analytical performance for the SARS-COV-2 N and S proteins

The SARS-CoV-2 N antigen immunoassay on the MSD platform had an average LOD of 0.45 pg/mL (0.25 to 0.93 pg/mL range), an LLOQ of 3.2 pg/mL, and a ULOQ of 50 ng/mL. The SARS-CoV-2 S antigen immunoassay had an average LOD of 6.2 pg/mL (2.1 to 9.0 pg/mL range), an LLOQ of 80 pg/mL, and a ULOQ of 250 ng/mL. Both assays were nonreactive or below detection limits for panel diluents, transport media, and common human coronavirus lysates OC43 and 229E.

### Distribution of N and S antigens in clinical samples

All 200 presumed positive samples and 100 of the 205 negative samples were analyzed by qRT-PCR for SARS-COV-2. Here, 182 samples were confirmed positive by qRT-PCR and 99 of the negatives were confirmed negative, with one repeatedly testing positive by qRT-PCR. The remaining negatives were assigned as qRT-PCR negative based on previously assigned qRT-PCR results, resulting in 183 PCR-positive samples and 222 PCR-negative samples. N and S antigen quantification was conducted on all 405 specimens. A positive correlation was found between the N antigen concentration and GEs (Figure 2A and 2B). The N antigen assays showed a high percent positive agreement (> 95%) for specimens containing 10^4^ or more GEs/mL SARS-CoV-2 (Figure 3). The percent positive agreement progressively dropped with decreasing concentrations of GEs. For the S antigen, the percent positive agreement was 86.4% at 10^5^ GEs/mL, with a sharp drop in agreement for lower concentrations. For all samples for which N antigen was within the LOQ of the assay (n = 120), the mean and median per GE N antigen observed were 12.7 fg and 1.5 fg N antigen (range of 0.1 to 204.2 fg/GE). For concentrations of SARS-CoV-2 GEs less than 10^4^ GEs/mL, the quantity of N antigen varied more widely for positive samples within the LOQ, with a trend toward highest N antigen per GE. For all samples for which S antigen was within the LOQ of the assay (n = 41), the mean and median per GE S antigen observed were 0.2 fg and 0.1 fg S antigen (range of 0.04 to 2.3 fg/GE).

**Figure 2.**
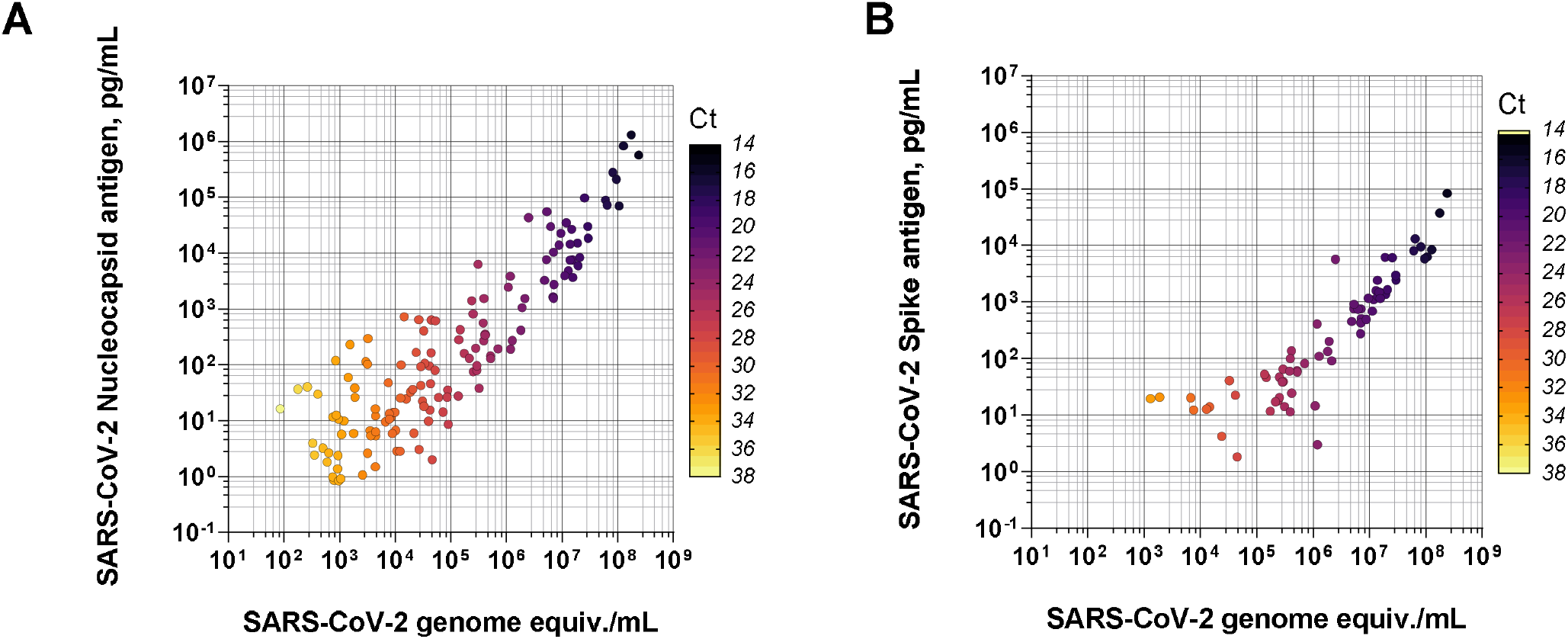
A. Correlation between N antigen concentration (pg/mL) and N genome equivalents (copies/mL). Mean cycle threshold (Ct) values are color coded to provide an indication of the corresponding Ct values. **B**. Correlation between S antigen concentration (pg/mL) and N genome equivalents (copies/mL). Mean Ct values are color coded to provide an indication of the corresponding Ct values.

**Figure 3.**
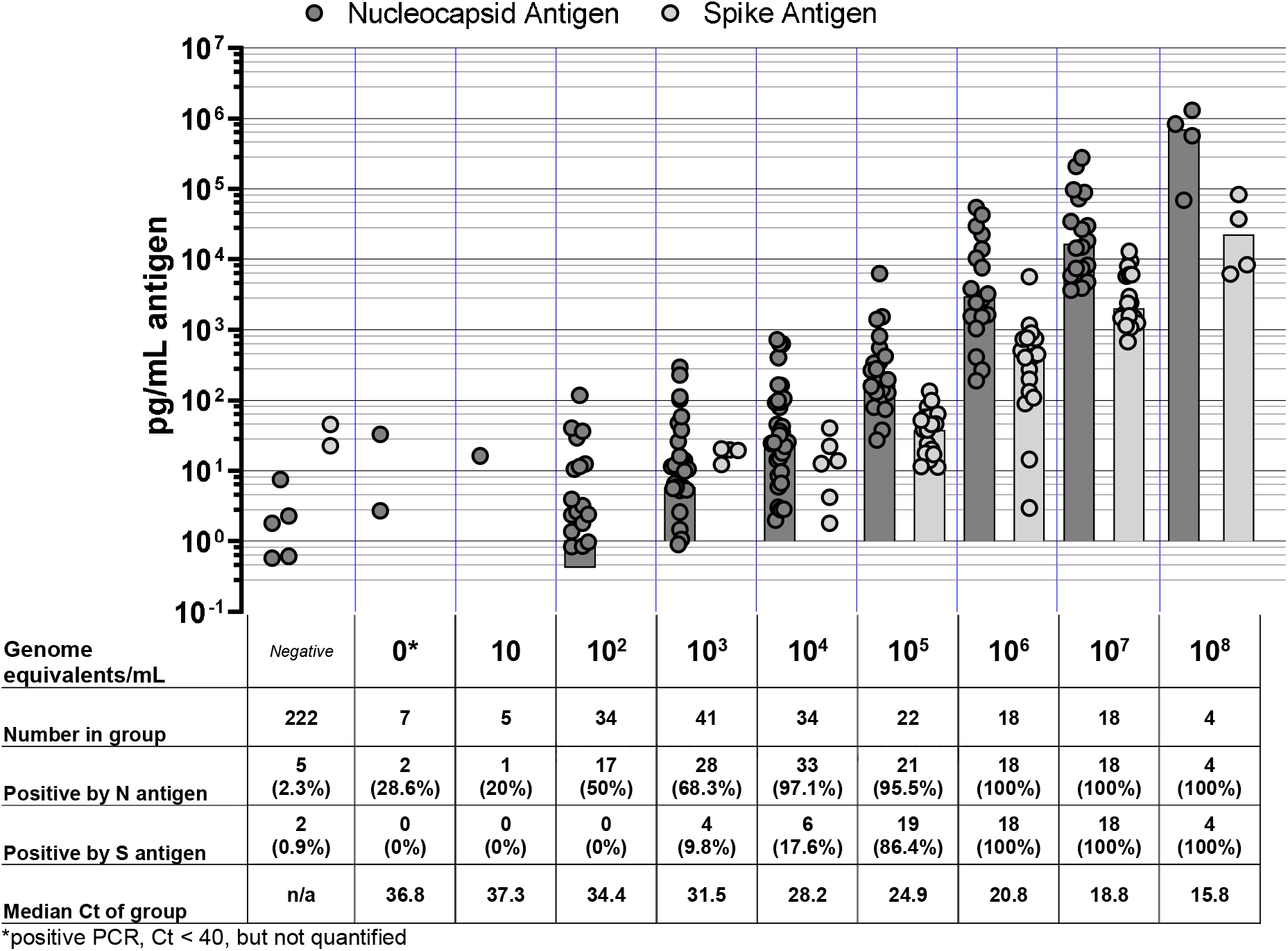
Antigen concentration over a range of SARS-CoV-2 genome equivalents detected by Meso Scale Discovery antigen quantification. Characteristics of groups of viral genome equivalent concentrations are shown in the table below the graph. Bars indicate the median of the group of antigen-positive points shown. Abbreviations: Ct, cycle threshold; N, nucleocapsid; PCR, polymerase chain reaction; S, spike.

### Benchmarking panels

Benchmarking panels were prepared to span concentrations of N antigen corresponding to LODs expected in rapid tests following dilution into the extraction buffer. As most RDTs only detect the N antigen, S antigen analysis was not included in the panel. HEK293-expressed and *E. coli*– expressed recombinant proteins were prepared in both buffer and negative swab pool dilution matrices, and linear fit of measured concentration as compared to target concentration was greater than 0.98 over the range of the panels of 0.2 ng/mL to 50 ng/mL.

### Quantification of inactivated virus and clinical dilutions, compared to genome equivalents

Though all panels were prepared from a single lot of inactivated SARS-CV-2 (BEI Resources), comparisons between two lots showed that for each lot, the per-calculated GE concentration of N antigen had a median of 1.0 fg/GE (range of 0.51 to 1.1 fg/GE) over the dilutions. In contrast, when compared to TCID_50_, the median amount of N antigen per TCID_50_ in the dilutions was 5,820 fg/TCID_50_ for BEI Resources lot 70033322 and 668 fg/TCID_50_ for lot 70035888. Both irradiated virus and clinical pool samples behaved similarly, even in terms of the per GE concentration of N antigen (Figure 4). For the clinical specimen pool dilution series, the median per GE quantity of N antigen was 2.4 fg/GE (range of 1.2 to 5.3 fg/GE).

**Figure 4.**
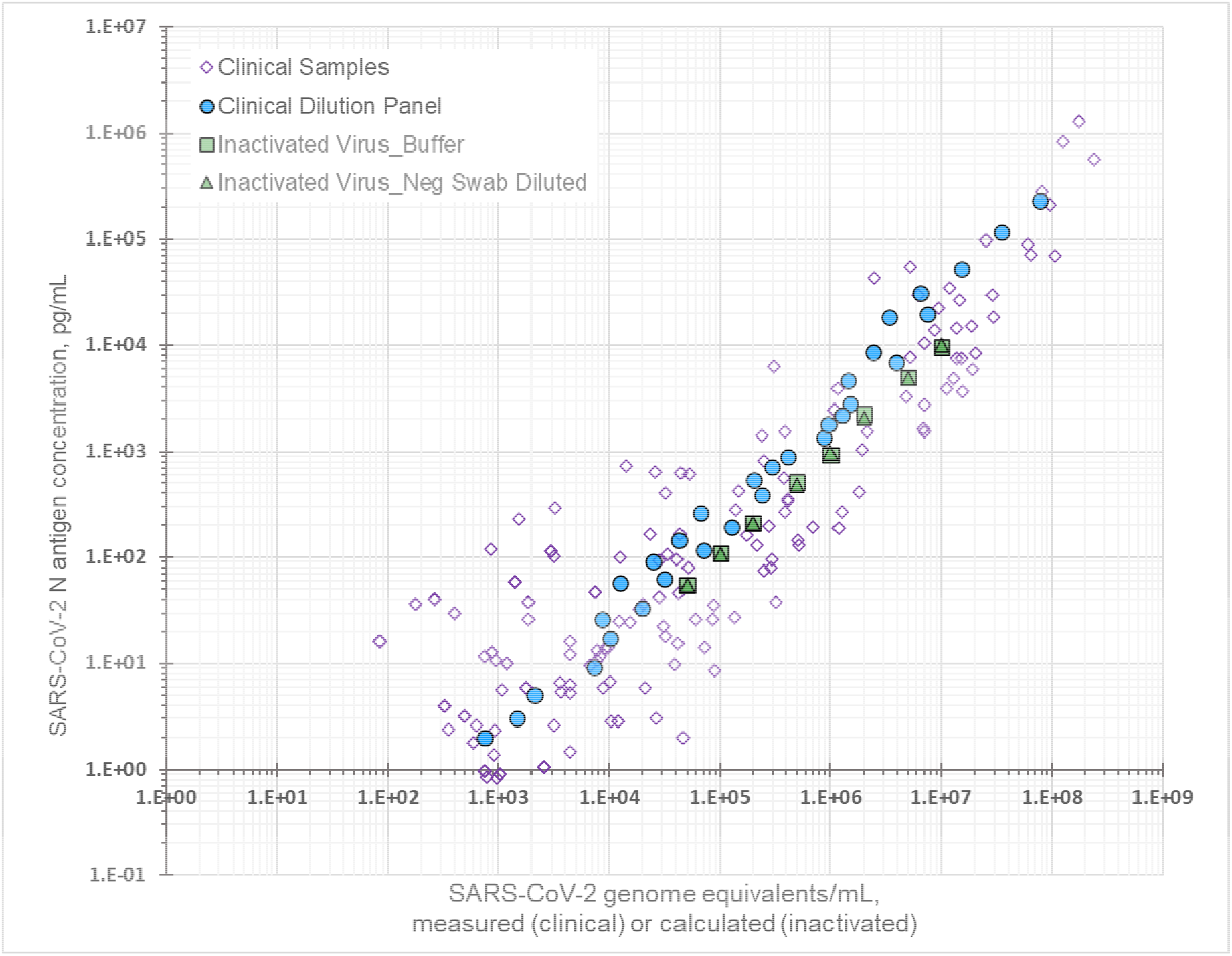
The relationhsip between N antigen concentration and genome equivalents for panels of diluted clinical specimens and inactivated virus. Blue circles indicate the clinical pool dilution series. Green squares and triangles indicate BEI Resources irradiated virus lot 7003588 in buffer and negative swab pool, respectively.

### Benchmarking of rapid antigen detection tests for SARS-CoV-2

Four RDTs were evaluated with the benchmarking panels: SD BIOSENSOR Abbott BinaxNOW COVID-19 Ag card test, Abbott Panbio COVID-19 Ag Rapid Test Device, LumiraDx SARS-CoV-2 Ag Test, and SD Biosensor STANDARD Q COVID-19 Ag Home Test. The tests were anonymized using identification numbers RDT 1 through RDT 4, in no particular order, for comparative presentations of the results.

### Reactivity of panel types

RDT line intensity scored either with a score card provided by the manufacturer or a preset universal score card allowed an assessment of the signal-dose response per panel member type. An illustrative example is given in Figure 5 for RDT 1. Recombinant proteins from mammalian and *E. coli*–based expression systems produced a strong dose-response signal on the RDTs and were similarly reactive. Greater reactivity was observed with both inactivated viral culture and diluted clinical positives, based on the final concentration of N antigen. Binary positive/negative results were used to estimate the probability of detection at different antigen concentrations. This was performed for all four tests (Figure 6). All tests performed best against the clinical specimen pool dilutions in terms of 90% probability of detection, and then differentially against the different panel members. RDTs had lower reactivity to the inactivated virus when it was diluted into the buffer diluent versus negative swab pool diluent (Table 2).

**Table 2.** Ninety percent probability of detection (95% confidence interval) of N antigen final concentration for benchmarking panel categories.

|  |  |  |  |  |  |
| --- | --- | --- | --- | --- | --- |
|  | Concentration of N antigen (final concentration added to test) with 90% probability of detection, pg/mL |  |  |  |  |
| Panel category ► | Clinical positive | Inactivated virus | Inactivated virus | Recombinant protein (*E. coli* and | Specificity and diluent controls |

| RDT number<br>▼ | Diluted in<br>negative swab<br>pool | Diluted in<br>negative swab<br>pool | Diluted in<br>buffer | mammalian<br>expressed)<br>Diluted in<br>buffer |  |
| --- | --- | --- | --- | --- | --- |
| RDT 1 | 46.774<br>(33.113–<br>60.256) | 93.325<br>(75.858–<br>114.815) | 125.893<br>(102.329–<br>151.356) | 181.97<br>(154.882–<br>208.93) | Not detected |
| RDT 2 | 23.988<br>(17.378–<br>31.623) | 54.954<br>(40.738–<br>72.444) | 58.884<br>(42.658–<br>79.433) | 74.131<br>(58.884–<br>91.201) | Not detected |
| RDT 3 | 102.329<br>(70.795–<br>141.254) | 144.544<br>(104.713–<br>194.984) | 229.087<br>(165.959–<br>301.995) | 208.93<br>(165.959–<br>263.027) | Not detected |
| RDT 4 | 21.38<br>(12.883–<br>32.359) | 22.909<br>(15.136–<br>33.884) | 42.658<br>(28.84–<br>63.096) | 128.825<br>(93.325–<br>181.97) | Not detected |

Abbreviations: *E. coli*, *Escherichia coli*; RDT, rapid diagnostic test.

**Figure 5.**
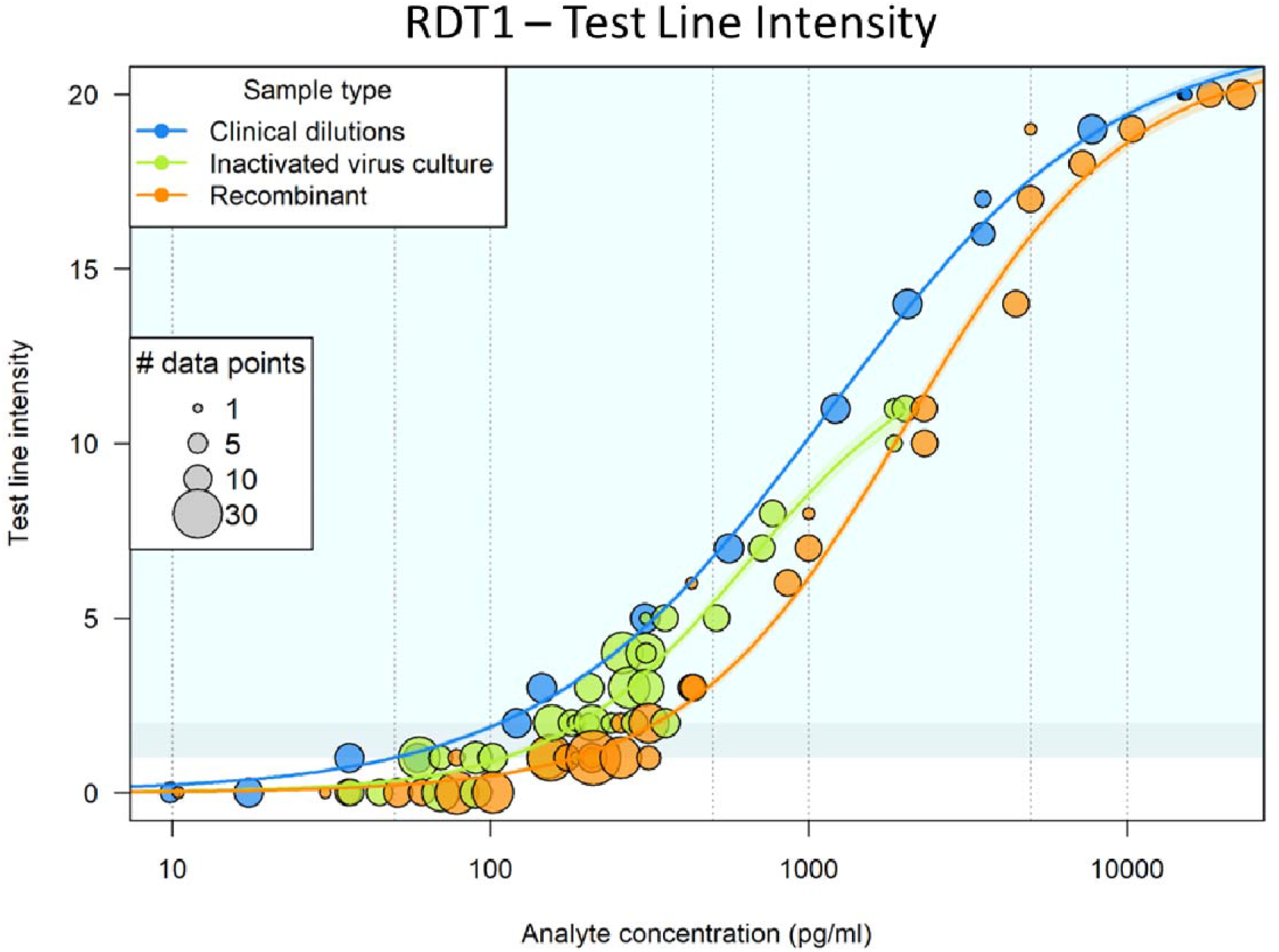
Illustrative sub-benchmarking panel member analysis for SARS-CoV-2 rapid diagnostic test 1. Circled positions indicate the replicates with a given test line intensity result for the concentration of antigen (analyte) panel added. Each panel subset has been given a different color code: blue for clinical specimen pool, green for inactivated virus, and orange for recombinant antigen. The size of the circles indicates the number of replicates supporting each data point. Test line intensity is shown on a scale of 0 (negative) and 1 through 20, representing from least to most intense visible test line of positive results. Abbreviation: RDT, rapid diagnostic test.

**Figure 6.**
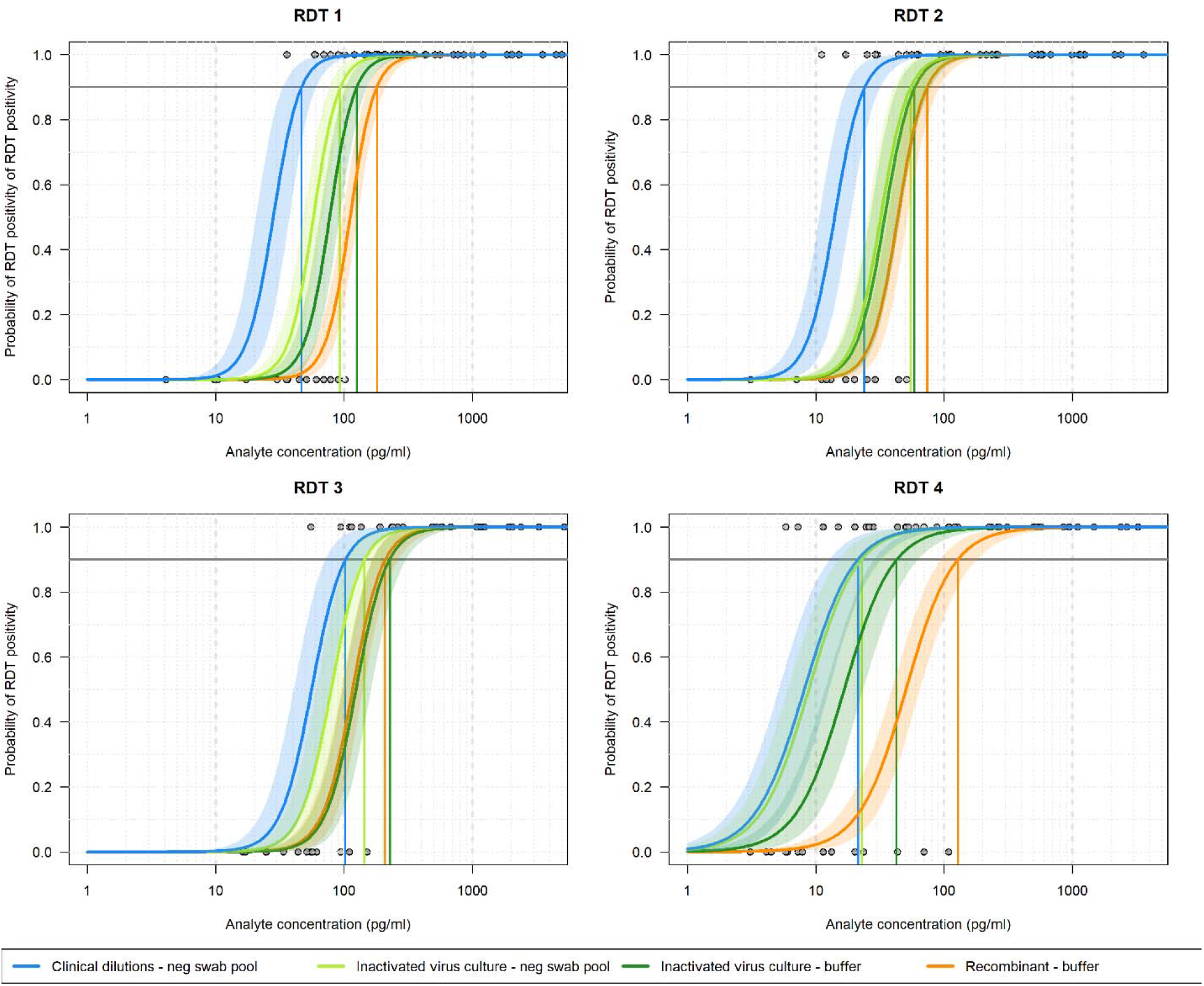
Probability of detection against antigen concentrations for the four antigen detection rapid diagnostic tests per benchmarking panel type and diluent matrix. Probability of detection of positive for the different benchmarking panel member types for each test in each panel (A, B, C, and D). Blue lines indicate clinical specimen dilution; green lines represent inactivated virus; and orange lines represent recombinant protein. Confidence intervals (95%) are indicated in matched-color shading. Abbreviation: RDT, rapid diagnostic test.

### Comparative benchmarking results

The clinical specimen pool dilution panels were plotted for all four tests, for comparison of analytical performance of the RDTs (Figure 7). The modeled 90% probabilities of detection were found to be 47, 24, 102, and 21 pg/mL of final concentration of N antigen added to test, for RDTs 1 through 4 respectively (Table 2).

**Figure 7.**
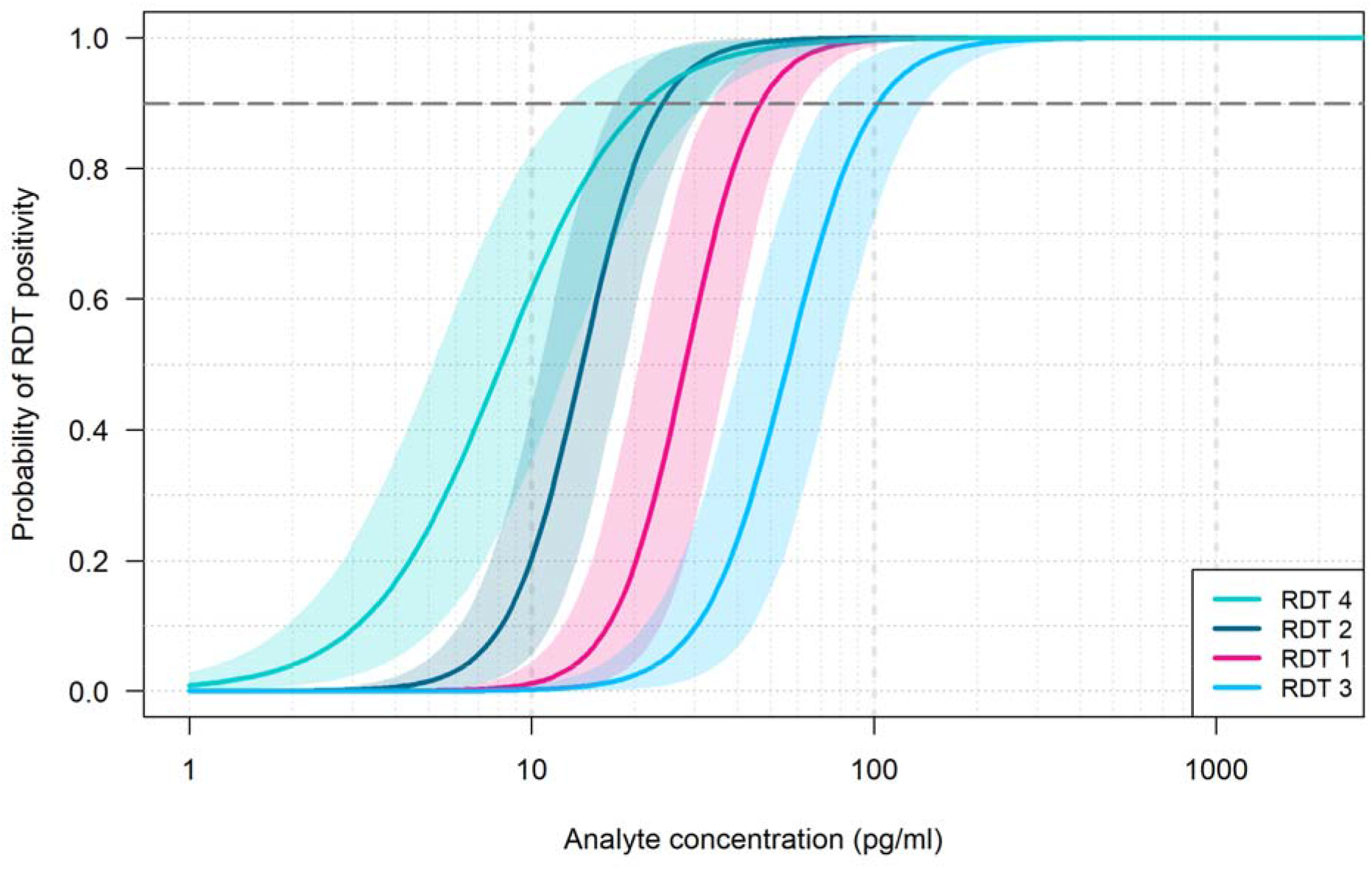
Probability of test positivity versus final N antigen (analyte) concentration added to test for clinical positive dilutions. The four curves indicate the probability of test positivity for each rapid antigen test product. The shaded lines indicate 95% confidence intervals. Abbreviation: RDT, rapid diagnostic test.

### Simulating clinical performance

One limitation of benchmarking is that the results are interpretable as a concentration inherently dependent upon assay configuration and the input volumes of the analyte. Interpretation of the final concentration of incoming analyte, as diluted into the assay extraction buffer, allowed normalization across RDTs but created the challenge of direct comparison to qRT-PCR values. If an identical swab were diluted into transport media (typically around 3 mL) and into RDT extraction buffer (typically around 300 µL), the volume difference could create a 10-fold disparity in analyte concentration from the swab. Figure 8 models this potential dilutional gain when using RDTs to predict what may be observed in a paired swab sampling. Limits of detection determined by the 90% cutoff were compared against the clinical sample set antigen concentrations, with the assumption that extracted material present in the PCR would instead be fully present in the extraction buffer for the rapid test, thus simulating a paired swab experiment. The detection limits based on the concentration of N antigen added to the test were found to affect the number of samples predicted to be detectable.

**Figure 8.**
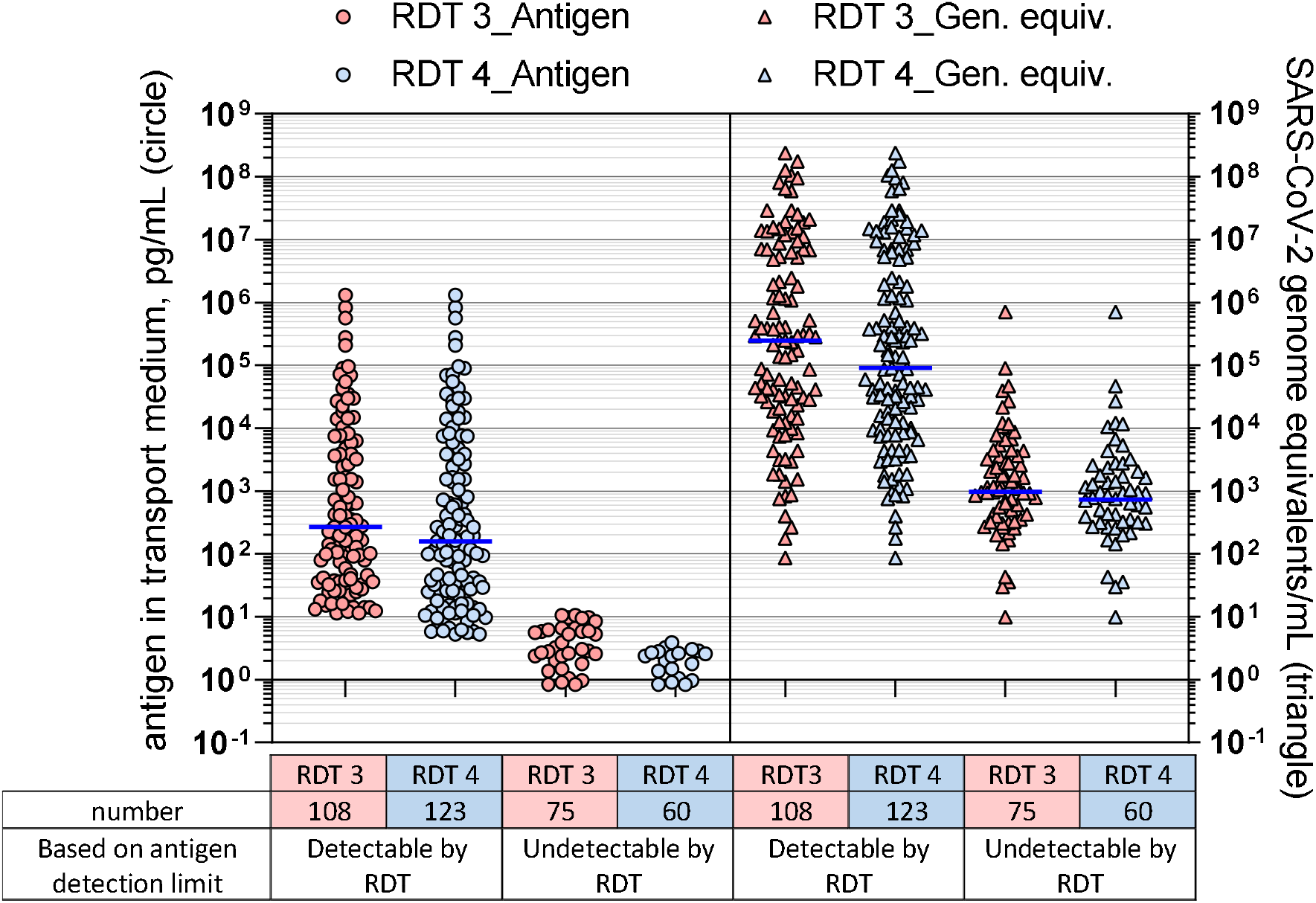
The predicted rapid diagnostic test (RDT) positivity from clinical samples based on their antigen detection limit. Results are for RDT 3 (pink shading) and RDT 4 (blue shading), shown as antigen concentration versus positivity (circle) or viral genome equivalent/mL versus positivity (triangle). Abbreviation: RDT, rapid diagnostic test.

RDTs 3 and 4, which had the highest and lowest analytical detection limits, respectively, based on final concentration, were compared in this simulation. Overlap in N antigen–detectable and – undetectable concentrations of GEs (Figure 8) was observed due to the spread of antigen concentration per GE relationship, but the median GEs/mL between the detectable and undetectable differed by about 2 orders of magnitude for all RDTs.

## DISCUSSION

An open platform assay was developed and described to quantify the N and S antigens in SARS-CoV-2–infected clinical specimens on the MSD platform. The assays showed good performance against RT-PCR–confirmed cases and a panel of negative specimens. Quantification of both the N and S antigens in specimens with associated viral load values showed a positive but not perfect correlation. As anticipated, a higher N antigen concentration was observed per GE in comparison to S antigen concentration, which has also been shown in plasma.^17,18^ These results support the focus on N antigen for RDTs and overall correlation of antigen concentration with genome copy number^9,19,20^.

The antigen assay combined with the qRT-PCR was used to pedigree a panel of reagents designed to benchmark N antigen RDTs. The benchmarking panel consisted of a dilution series of recombinant N antigens, expressed in both prokaryotic and eukaryotic expression systems, two sources of inactivated virus, and a clinical specimen pool. Four widely used tests, either cleared by EUL or EUA, were evaluated against the panel: SD Biosensor STANDARD Q COVID-19 Ag Home Test, Abbott COVID-19 Ag Rapid Test Device, Abbott BinaxNOW COVID-19 Ag card test, and LumiraDx SARS-CoV-2 Ag test.

*E. coli*–expressed recombinant N antigen behaved similarly to N antigen expressed in mammalian cells on the N antigen assay. Inactivated virus contained N antigen concentration per viral GEs within the same range as observed in clinical samples. Comparison of the relationship between N antigen concentration and GEs versus TCID_50_ across two lots from BEI Resources suggests that GEs are more reflective of the anticipated N antigen concentration.

The benchmarking results for the four tests highlight a range of reactivities against the different panel components. Consistently, all tests showed improved LOD to the clinical specimens over the other N antigen sources (inactivated virus and recombinant antigen). RDT 4 showed an improved response to negative swab diluent over buffer and highlights the need for comprehensive and test-specific data with a variety of materials before conclusions can be made. Understanding the reactivity against inactivated virus and recombinant protein is valuable, as these can be readily expressed, noninfectious sources of N antigen for emerging virus strains for which there may be a concern for sequence-dependent false negativity.^21–24^

The 90% probability of detection, a proxy for LOD, for the clinical specimen pool ranged from 20 pg/mL to 100 pg/mL across the four tests. While the 90% probability of detection limit model is not a standardized method, it allows a continuous detection response function to be fit to standardized panels, rather than requiring custom dilutions for each test.^25^ The simulated paired swab results generated by applying analytical detection limits to clinical samples was in alignment with observed clinical performance evaluations, indicating that RDTs detect a high percentage of infections with viral loads associated with Ct values less than 30.^26,27^

While the analytical performance measured through benchmarking may be strongly indicative of the clinical performance of the tests, it cannot be correlated directly to final performance. Some of the factors that will influence the final performance are (1) relative efficiencies for absorption and release of N antigen by the manufacturer’s specific nasal swab and elution buffer; (2) the dilution factor and original specimen equivalents loaded on to the test after all processing is conducted; and (3) differing sensitivities of the test to the circulating SARS-CoV-2 strains in the population being sampled. Although RDTs showed the best analytical performance against the diluted clinical specimens pool, inclusion of recombinant protein sources can readily incorporate N antigens into the benchmarking panel with nonsynonymous mutations that may alter the analytical performance of the diagnostic test.^21,22,28^

## CONCLUSION

The benchmarking panel allowed rapid assessment of the analytical performance by manufacturers and third parties in a manner that could be directly compared across RDTs for SARS-CoV-2. Full characterization of both molecular and protein analytes allowed for comparison of results across testing platforms. The benchmarking results were complementary to efforts to produce international standards and to support clinical evaluations.

## Data Availability

All data produced in the present study are available upon reasonable request to the authors

## ABBREVIATIONS

BEI: BEI Resources
BSA: bovine serum albumin
CDC: US Centers for Disease Control and Prevention
COVID-19: coronavirus disease 2019
Ct: cycle threshold
*E. coli*: *Escherichia coli*
EUA: emergency use authorization
EUL: emergency use license
GE: genome equivalent
LOD: limit of detection
LLOQ: lower limit of quantification
MSD: Meso Scale Discovery
N: nucleocapsid
NIAID: US National Institute of Allergy and Infectious Diseases
NIH: US National Institutes of Health
PBS: phosphate-buffered saline
qRT-PCR: quantitative reverse transcription polymerase chain reaction
RDT: rapid diagnostic test
RT-PCR: reverse transcription polymerase chain reaction
S: spike
SARS-CoV-2: severe acute respiratory syndrome coronavirus 2
TCID_50_: 50% tissue culture infective dose
ULOQ: upper limit of quantification.

## ACKNOWLEDGMENTS

The authors would like to acknowledge funding by the Bill&Melinda Gates Foundation (https://www.gatesfoundation.org/) via grant INV-016821. The funder did not have any role in the study design, data collection and analysis, decision to publish, or preparation of the manuscript. Laboratories providing samples used in this work, accessed through the Washington COVID-19 Biorepository, were The Everett Clinic (part of Optum), FidaLab, Northwest Pathology, Bloodworks Northwest, the Washington State Public Health Laboratories, and the University of Washington School of Medicine. Rapid diagnostic tests used in this work were either purchased using grant funding from the Bill&Melinda Gates Foundation, or donated by partners in support of research via grant from Abbott Laboratories and agreements from LumiraDx and the Washington State Department of Health. Donors did not have any additional role in the study design, data collection and analysis, decision to publish, or preparation of the manuscript. Finally the authors would like to thank Brook Alemayehu and Terri Scott for editorial support with the manuscript.

